# Emergence of the Novel Sixth *Candida auris* Clade VI in Bangladesh

**DOI:** 10.1101/2024.04.12.24305665

**Authors:** Tahsin Khan, Naimul Islam Faysal, Md Mobarok Hossain, Syeda Mah-E-Muneer, Arefeen Haider, Shovan Basak Moon, Debashis Sen, Dilruba Ahmed, Lindsay A. Parnell, Mohammad Jubair, Nancy A. Chow, Fahmida Chowdhury, Mustafizur Rahman

**Affiliations:** Genome Centre, Infectious Diseases Division, icddr,b, Mohakhali, Dhaka-1212, Bangladesh; Emerging Infections, Infectious Diseases Division, icddr,b, Mohakhali, Dhaka-1212, Bangladesh; Mycology Laboratory, icddr,b, Mohakhali, Dhaka-1212, Bangladesh; Mycotic Diseases Branch, Centers for Disease Control and Prevention, Atlanta, Georgia, USA

## Abstract

*Candida auris*, initially identified in 2009, has rapidly become a critical concern due to its antifungal resistance and significant mortality rates in healthcare-associated outbreaks. To date, whole-genome sequencing (WGS) has identified five unique clades of *C. auris*, with some strains displaying resistance to all primary antifungal drug classes. In this study, we presented the first WGS analysis of *C. auris* from Bangladesh, describing its origins, transmission dynamics, and antifungal susceptibility testing (AFST) profile. Ten *C. auris* isolates collected from hospital settings in Bangladesh were initially identified by CHROMagar™ Candida Plus, followed by VITEK^®^2 system and later sequenced using Illumina NextSeq 550 system. Reference-based phylogenetic analysis and variant calling pipelines were used to classify the isolates in different clades. All isolates aligned ∼90% with the Clade I *C. auris* B11205 reference genome. Of the ten isolates, eight clustered with Clade I isolates, highlighting a South Asian lineage prevalent in Bangladesh. Remarkably, the remaining two isolates formed a distinct cluster, exhibiting >42,447 SNP differences compared to their closest Clade IV counterparts. This significant variation corroborates the emergence of a sixth clade (Clade VI) of *C. auris* in Bangladesh, with potential for international transmission. AFST results showed that 80% of the *C. auris* isolates were resistant to fluconazole and voriconazole, whereas Clade VI isolates were susceptible to azoles, echinocandins, and pyrimidine analogue. Genomic sequencing revealed *ERG11*_Y132F mutation conferring azole resistance while *FCY1*_S70R mutation found inconsequential in describing 5-flucytosine resistance. Our study underscores the pressing need for comprehensive genomic surveillance in Bangladesh to better understand the emergence, transmission dynamics, and resistance profiles of *C. auris* infections. Unveiling the discovery of a sixth clade (Clade VI) accentuates the indispensable role of advanced sequencing methodologies.

**IMPORTANCE:** *Candida auris* is a nosocomial fungal pathogen which is commonly misidentified as other *Candida* species. Since its emergence in 2009, this multi-drug resistant fungus has become one of the five urgent antimicrobial threat by 2019. Whole Genome Sequencing (WGS) has proven to be the most accurate identification technique of *C. auris* which also played a crucial role in the initial discovery of this pathogen. WGS analysis of *C. auris* has revealed five distinct clades where isolates of each clade differ among themselves based on pathogenicity, colonization, infection mechanism as well as other phenotypic characteristics. In Bangladesh, *C. auris* was 1^st^ reported in 2019 from clinical samples of a large hospital of Dhaka city. To understand the origin, transmission dynamics and antifungal resistance profile of *C. auris* isolates circulating in Bangladesh, we conducted WGS based surveillance study on two of the largest hospital settings in Dhaka, Bangladesh.

## INTRODUCTION

*Candida auris*, an emerging multidrug-resistant pathogen, imposes a serious threat to global public health owing to its high virulence and transmissibility in hospital settings [1]. Since the first report in 2009 in Japan, this nosocomial pathogen has already been reported in over 50 countries, colonizes human skin and hospital environments, and causes invasive candidiasis, especially in critically ill and immunosuppressed patients [2–4]. Patients with *C. auris* infection or colonization frequently exhibit other underlying health conditions and comorbidities, such as diabetes, cardiovascular diseases, liver diseases, chronic or acute kidney failure, solid tumor or malignancies [5].

Whole-genome sequencing (WGS) and matrix-assisted laser desorption ionization–time of flight (MALDI-TOF MS) platforms are the best-suited options to correctly identify *C. auris* [6]. Worldwide, WGS analysis has grouped *C. auris* into five distinct genotypic clades based on 40,000 to >200,000 single nucleotide polymorphism (SNPs) such as Clade I (South Asia), Clade II (East Asia), Clade III (Africa), Clade IV (South America), and Clade V (Iran) [3, 7, 8]. Though the reasons for the reported independent and simultaneous emergence in different geographical regions are unknown, animal reservoirs and environmental changes are speculated to accelerate this situation [9]. Each clade harbors highly related isolates due to local transmission and clonal expansion within countries [10, 11]. However, analysis from molecular epidemiological investigations, outbreaks, and individual cases also revealed genetic complexity suggesting multiple introductions followed by local transmissions in different countries [12].

Antifungal resistance seems to be clade specific [13]. Clade II isolates are generally more susceptible to antifungal agents, whereas Clade I, III and IV are highly resistant [1, 8, 14]. Only 2/5 identified isolates of Clade V were found resistant to fluconazole but were susceptible to triazoles, echinocandins, and amphotericin B [15, 16]. Nonetheless, antifungal resistances were observed on all five clades to currently used antifungal drugs such as azoles (fluconazole, voriconazole, itraconazole, and isavuconazole), polyenes (amphotericin B), echinocandins (caspofungin, micafungin, and anidulafungin), and pyrimidine analogue flucytosine [11, 17–19]. Genomic level inspections revealed clade-specific azole-resistant mutations in *ERG11*: Y132F or K143R in Clade I, F126T/L in Clade III, Y132F in Clade IV, and Clade V [3, 7]. Numerous mechanisms responsible for antifungal resistance have been reported for *C. auris*, such as echinocandin-resistant *FKS1*:S639/F/P/Y, and *FKS1*:S635Y/P mutations, 5-flucytosine resistant *FCY1*:S70R, *FCY2*:M128fs (frameshift), *ADE17*:G45V and *FUR1*:1Δ33 [20, 21]. Resistance mechanisms for amphotericin B are not well documented; however, WGS analysis has identified several nonsynonymous polymorphisms that might infer resistance [22].

Bangladesh (BD) is located in South Asia where fungal outbreaks have been widely reported [23–25]. The first reported case of *C. auris* in Bangladesh was in 2019, whereas India and Pakistan reported in 2013 and 2015 respectively [7, 26, 27]. Dutta et al., 2019 reported *C. auris* infections in 19 patients that showed resistance to voriconazole (18/21), fluconazole (14/21), and amphotericin B (5/21) [28]. A recent study that conducted AFST for major fungal pathogens in different hospital settings in Bangladesh also reported *C. auris* resistance against fluconazole (100%), itraconazole (100%), voriconazole (33.3%), and amphotericin B (100%) [29]. Monitoring the circulating clades and observing the resistance pattern of *C. auris* is crucial to prevent spread and transmission. Considering the absence of genomic surveillance data for *C. auris* in Bangladesh, we conducted WGS analysis on 10 *C. auris* isolates collected from intensive Care unit (ICU) and neonatal intensive care unit (NICU) patients from two national hospitals in Bangladesh. We report the emergence of a sixth clade (Clade VI), circulating along with Clade I isolates in Bangladesh.

## RESULTS

### Patient Information

Ten *C. auris* isolates representing eight ICU patients and two NICU patients were described in this study. Patients were admitted with various underlying health complications; none had candidemia (**File S1**). Among the patients, only one patient (101022) from ICU had previous travel history to the United Arab Emirates within one year of enrolment. Overall, 40% (4 patients from ICU) developed nosocomial colonization with *C. auris*, and none received any antifungal treatment since patients either expired or were discharged by the time the test result was released (Table 1).

**TABLE 1:**
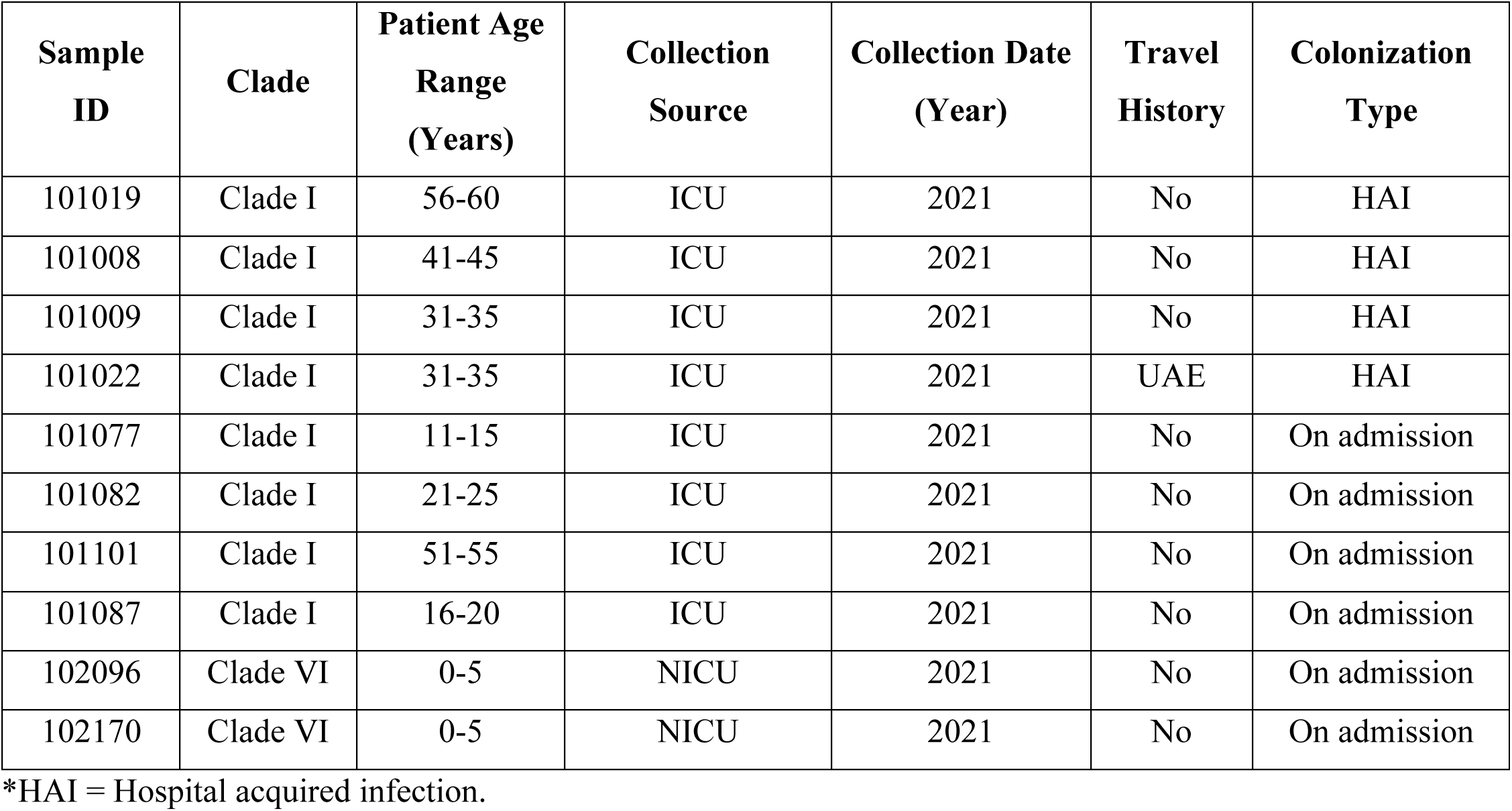
Patient metadata of sequenced *Candida auris* isolates.

### WGS SNP Calling and Phylogeny Analysis: Emergence of a Novel Clade

Whole genome-based SNP calling was performed to distinguish all BD isolates in different clades, incorporating a total of 25 *C. auris* isolates that represent the known five clades (**File S2**). Around 92.68% (SD ±1.17) reads of BD isolates mapped with the reference B11205 with an average GC content of 44.41% (SD ±0.15) (**File S3**). Phylogenetic analysis revealed that BD-ICU isolates (Sample ID 101019, 101008, 101009, 101022, 101077, 101082, 101101, and 101087) clustered closely with Clade I with a maximum SNP difference of 163 SNPs (Fig. 1, **File S4**). Notably, these isolates differed by only ≤17 SNPs from each other except isolates 101019 and 101022. The isolate (101022) collected from the ICU patient who traveled to UAE was separated by only 18 SNPs from the UAE_B14000 isolate. Isolate 101019 had the highest SNP differences with other Clade I BD-ICU isolates (87-102 SNPs). Within the Clade I clusters, Clade I BD isolates had closer resemblance to UK_15B5 isolate (51-86 SNPs) than those from India (108-155 SNPs), Pakistan (109-148 SNPs) and New Jersey, USA (124-163). Interestingly, the BD-NICU isolates (102096, and 102170) clustered separately from all known clade isolates (Fig. 1). These two isolates differed by only 68 SNPs and were genetically distinct from all the clades by at least 42,447 single-nucleotide polymorphisms, and hence, formed a novel clade, Clade VI [7]. South American Clade (Clade IV) was the closest neighbor (>42,447 SNPs) for Clade VI, whereas the Iranian Clade (Clade V) was the most distant neighbor (>254,068 SNPs). To pinpoint the origin of this novel clade, we inquired the neonates’ parents about any international travel history, but none had any in 2020 or before. The two neonates were born in different hospitals, but later they were transferred to the NICU of study hospital. It was unknown whether their parents were suffering from fungal colonization or infections; or whether these neonates acquired *C. auris* colonization during birth or from the previous hospitals. To investigate the emergence of this novel clade, phenotypic differences between the Clade I and Clade VI isolates were observed and are described in Table 2 (**Fig S1)**.

**FIG 1.**
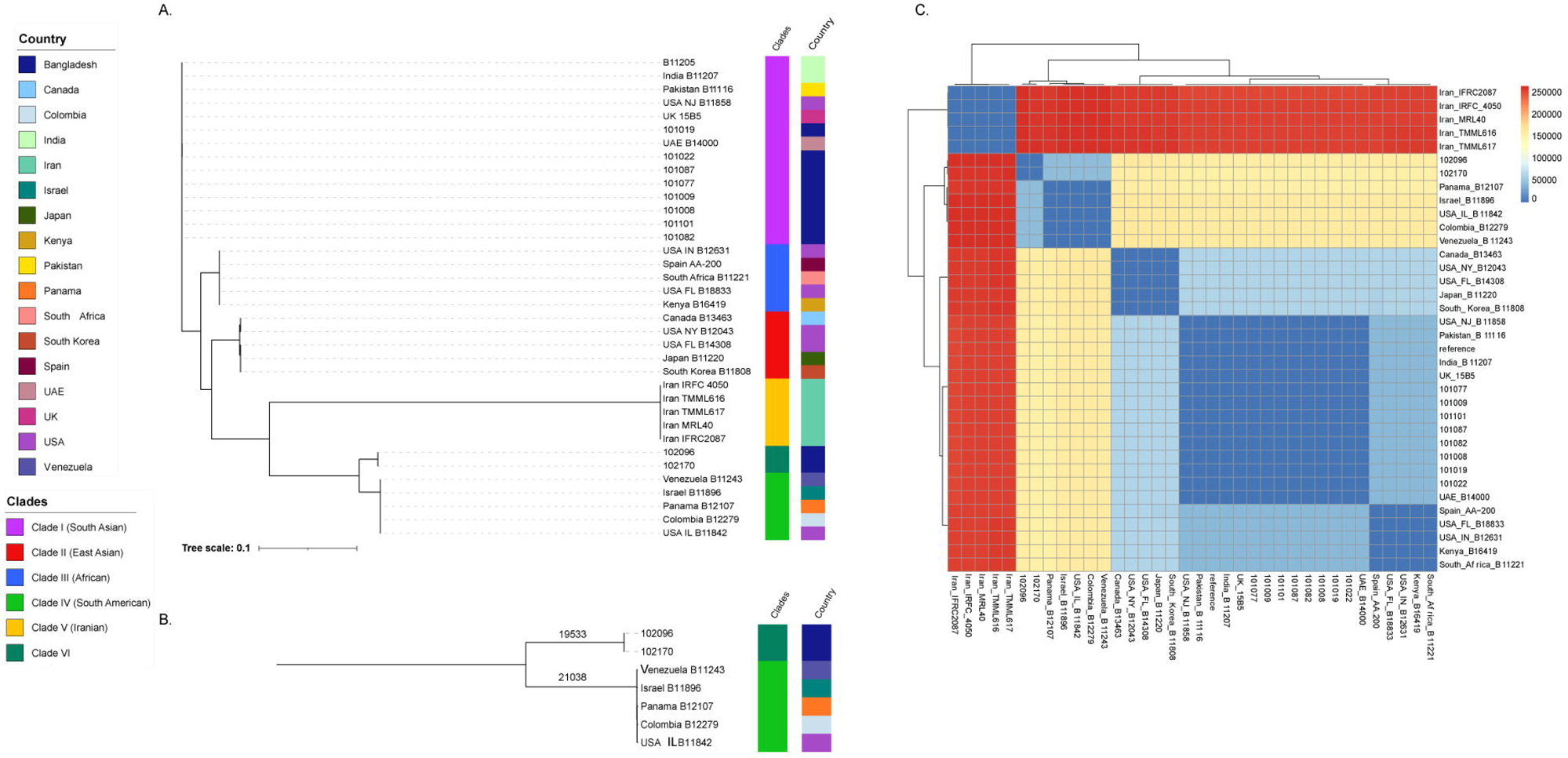
Phylogenetic tree based on SNPs of 35 *C. auris* isolates. (A) Representation of known five clades with novel Clade VI of *C. auris*. (B) Distances between Clade IV and novel Clade VI. All the BD isolates from our study are represented by dark blue color. Eight BD-ICU isolates clusters with reference isolate and 5 Clade I isolates from different countries. Two BD-NICU isolate (102096, and 102170) clusters together distinctly from other clades forming novel Clade VI. Isolate B11243 of Clade IV has the closest SNP distances from Clade VI isolates. (C) Clade-wise heatmap of SNP distances among all *C. auris* isolates used in this study.

**TABLE 2:**
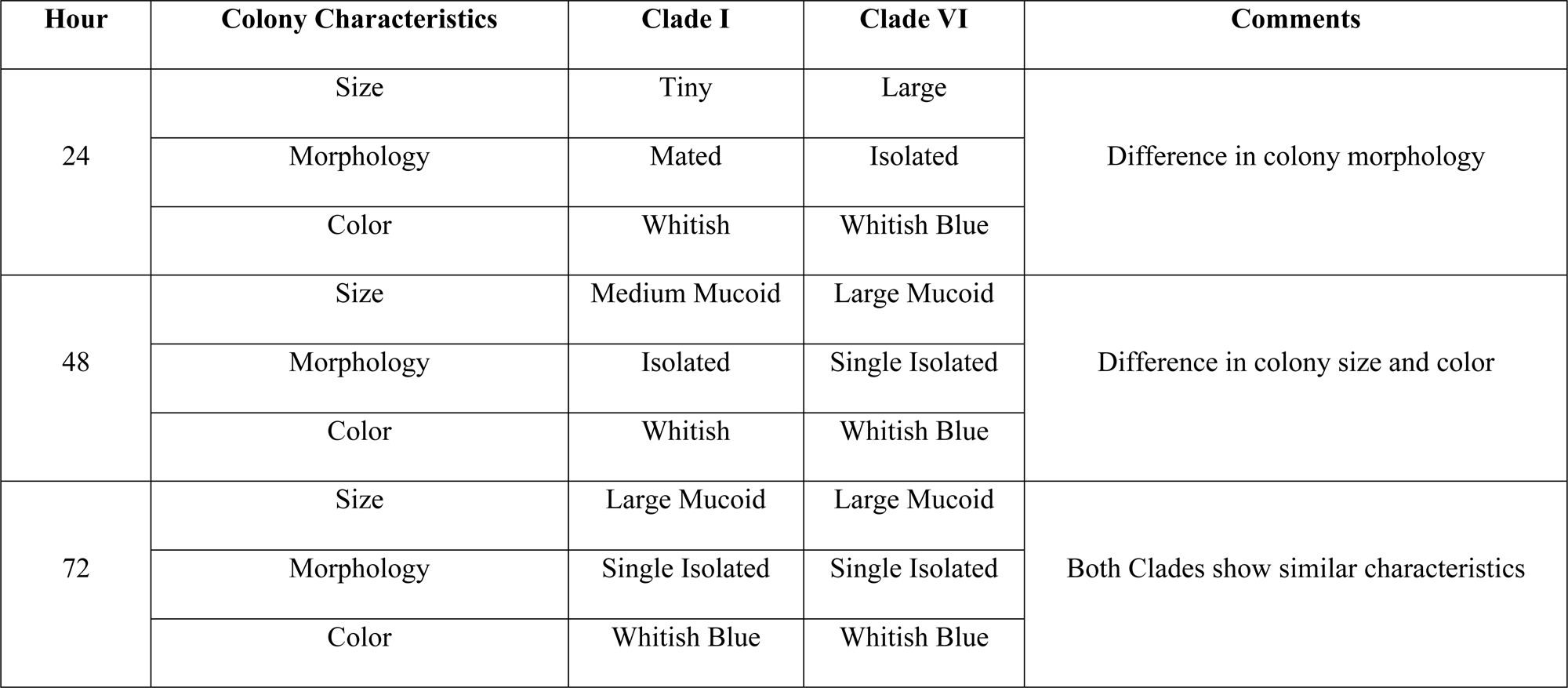
Phenotypic differences observed between BD Clade I and Clade VI isolates of *Candida auris* at different timelines (A) 24 hours (B) 48 hours and (C) 72 hours.

### Molecular Resistance Mechanisms

All the BD isolates were screened for mutations associated with antifungal resistance. For fluconazole and other triazole resistance, *ERG11, CDR1, TAC1B* and *MRR1A* gene mutations were screened in several positions leading to increased MIC values. All BD-ICU isolates of Clade I harbored *ERG11*_Y132F substitution. No mutations were detected in *CDR1, TAC1B* and *MRR1A* genes. No known *FKS1* gene mutation was observed in BD-ICU isolates which is reflected by susceptibility to caspofungin in AFST results (Table 3). *FCY1*_S70R mutation which confers 5-flucytocine resistance was detected in all BD-ICU isolates by genomic analysis. No known mutations in *ERG3* and *ERG6* genes were detected which could explain the amphotericin B resistance in isolate 101022. Surprisingly, Clade VI isolates from BD-NICU patients had no known mutations in any of the drug resistance genes (Table 3).

**TABLE 3:** Results of known mutation analysis and antifungal susceptibility test for azoles, polyenes, and echinocandins.

| Sample ID | Clade | Azole |  | Echinocandin | Polyene | Gene Mutations |  |
| --- | --- | --- | --- | --- | --- | --- | --- |
|  |  | Fluconazole | Voriconazole | Caspofungin | Amphotericin B | *ERG11* | *FCY1* |
| 101019 | Clade I | 256 | 32 | 0.19 | 1 | Y132F | S70R |
| 101008 | Clade I | 256 | 32 | 0.25 | 1 | Y132F | S70R |
| 101009 | Clade I | 256 | 32 | 0.38 | 1 | Y132F | S70R |
| 101022 | Clade I | 256 | 32 | 0.125 | 2 | Y132F | S70R |
| 101077 | Clade I | 256 | 32 | 0.25 | 0.5 | Y132F | S70R |
| 101082 | Clade I | 256 | 32 | 0.19 | 0.75 | Y132F | S70R |
| 101101 | Clade I | 256 | 32 | 0.25 | 0.5 | Y132F | S70R |
| 101087 | Clade I | 256 | 32 | 0.25 | 1 | Y132F | S70R |
| 102096 | Clade VI | 6 | 0.19 | 0.125 | 0.75 | - | - |
| 102170 | Clade VI | 8 | 0.19 | 0.125 | 2 | - | - |

### Antifungal Susceptibility Testing (AFST)

Antifungal susceptibility testing was performed on all BD *C. auris* isolates against fluconazole, voriconazole, caspofungin, and amphotericin B (Table 3). Based on U.S. CDC tentative breakpoints, all BD-ICU isolates were highly resistant to fluconazole (MIC 256 mg/L) and voriconazole (MIC 32 mg/L). Isolate 101022 collected from patient with recent travel history to the United Arab Emirates additionally showed resistance to amphotericin B (MIC 2 mg/L) along with fluconazole and voriconazole. No resistance was observed for caspofungin in BD-ICU isolates. On the contrary, BD-NICU isolates representing Clade VI were susceptible to 3 major antifungal drug classes. However, BD-NICU isolate 102170 demonstrated resistance to amphotericin B (MIC 2 mg/L). Only isolate 101019 of Clade I harboring *FCY1_*S70R mutation showed resistance against 5-flucytosine in both disk diffusion and automated VITEK^®^2 methods, while other Clade I isolates showed sensitivity in both methods even though these harbor the same mutation (Table 4). Mutational analysis on the resistance profile aligned with the AFST results except for isolate 101022 and 102170; no known mutation was found that confer amphotericin B resistance.

**TABLE 4:**
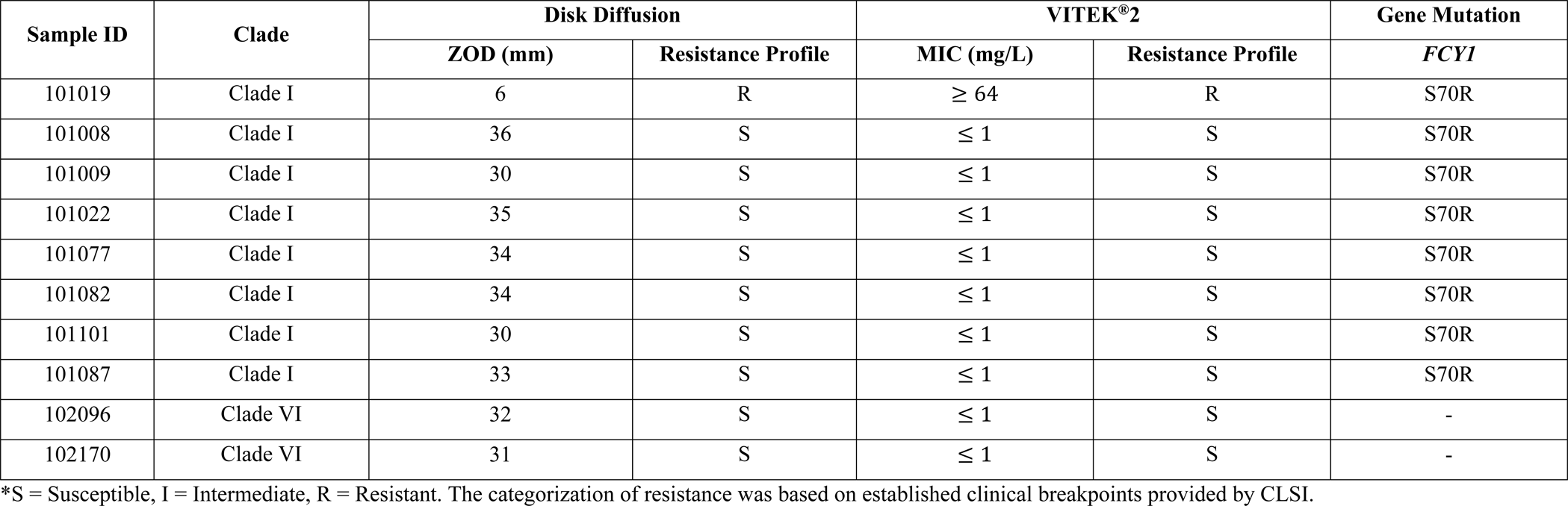
Results of 5-flucytosine antifungal susceptibility test by disk diffusion and VITEK^®^2 method.

## DISCUSSION

*Candida auris,* first detected in Bangladesh in 2019, has emerged as a global multi-drug resistant healthcare-associated pathogen [29]. WGS-based approach revealed the circulation of Clade I lineages, and the emergence of a new Clade VI in Bangladesh. Six BD-ICU Clade I isolates (101008, 101009, 101077, 101082, 101101, 101087) collected from adult patients, who never traveled overseas, are phylogenetically closest to each other (SNPs range 3-17), indicating the ongoing local and clonal transmission [12, 30]. The isolate 101022 from an ICU patient who previously traveled to UAE and died after admission was only 18 SNPs different than UAE_B14000, indicating possible Clade I transmission from UAE to Bangladesh.

The genome sequencing approach separated the clades of *C. auris* by tens of thousands of SNP differences [7]. The ∼42,447 SNPs difference of BD-NICU isolates (102096 and 102170) indicate the emergence of the new clade, Clade VI, in Bangladesh. Though tracing was not possible to identify the source of this nosocomial infection in neonates, it is evident that Clade VI is circulating in Bangladesh. By the time of writing this manuscript, a preprint published by a group of researchers from Singapore claimed the presence of a new clade separated by >36,000 SNPs from all known five clades of *C. auris* [31]. These isolates namely isolate A (LIMS ID F3485, Biosample SAMN36753178), Isolate B (LIMS ID F1580, Biosample SAMN36753179), Isolate C (LIMS ID F0083, Biosample SAMN36753180), and 102096 from our study (SRR24877248) were distinct by at least 70 SNPs [31]. Isolate C of the novel clade was collected from the blood of a Bangladeshi patient in 2018, who was airlifted directly to Singapore General Hospital to have treatment, and was initially classified as Clade IV [30]. The presence of candidemia in this patient indicates the undetected circulation of novel *C. auris* clade in Bangladesh and a later transfer to Singapore in 2018 before the first detection of *C. auris* in Bangladesh in 2019 [28]. It is surprising to note that even though Clade I and Clade VI isolates are circulating in Bangladesh simultaneously, Clade VI isolates acquired no known mutations and showed no phenotypic resistance except amphotericin B (Isolate 102170). It is of utmost importance to conduct nationwide genomic surveillance of prospectively collected *C. auris* isolates along with archived isolates stored at different hospitals in Bangladesh to understand the origin and transmission dynamics of Clade VI.

Clade I BD-ICU isolates showed the general trend of azole resistance for both mutational (genotypic) and AFST (phenotypic) results against fluconazole and voriconazole [7]. Surprisingly, these isolates showed inconsistency for 5-flucytosine resistance pattern except for isolate 101019. A similar observation was reported from a 29-year-old woman residing in New Jersey, who was diagnosed with severe short bowel syndrome [20]. She underwent a multi-visceral transplantation surgery and during her post-operative period of 72 days, the patient developed refractory fungal peritonitis. Nineteen *C. auris* isolates were obtained from the patient at various time intervals. While the initial 12 isolates collected within the initial 33 days demonstrated sensitivity to 5-flucytosine, the subsequent 7 isolates obtained between days 33 and 72 exhibited resistance to this antifungal agent. Genomic analysis of these isolates revealed the presence of the *FCY1*_S70R mutation in all 19 isolates. The authors argued *FCY1_*S70R mutation to be inconsequential in describing 5-flucytosine resistance. The findings in our study reflected the discrepancy between *FCY1_*S70R mutation and resistance pattern for Clade I isolates, strengthening the argument of inconsequential behavior of *FCY1_*S70R mutation in describing the 5-flucytosine resistance. Two BD-NICU isolates representing Clade VI exhibited no resistance to the antifungals tested (except amphotericin B
for Isolate 102170) and harbored no known mutations conferring resistance. Active clinical surveillance, monitoring of clinical symptoms, and extensive epidemiological investigation will accelerate understanding of the significance of this newly discovered sixth clade of *C. auris*.

## LIMITATIONS

Our study presents the WGS-based approach to detect the circulation of *C. auris* clades for the first time in Bangladesh. The isolates sequenced in this study were collected from the archived samples in 2021, as part of regular surveillance to monitor infections and colonization associated with ICU/NICU admitted patients, tracing the contacts of *C. auris* positive cases was not performed at that time. The small sample size in this study may not be the representative picture of *C. auris* circulation in Bangladesh. Including a large sample size would increase the chances of capturing other undetected clades circulating in Bangladesh. Long-read sequencing would ensure the recovery of the nearly full genome and the comprehensive genomic study on the antifungal resistance profile. Overall, our study demonstrates the necessity of *C. auris* genomic surveillance to understand the transmission dynamics, pathobiology and containment of this urgent antimicrobial-resistant threat in Bangladesh.

## CONCLUSION

We report the circulation of Clade I *C. auris* in Bangladesh along with the emergence of the novel Clade VI detected patients in the ICU and NICU. Bangladesh remains the hub of this novel clade since a Clade VI isolate was also found in the blood of a Bangladeshi patient airlifted in Singapore in 2018. It is critical that we enforce nation-wide genomic surveillance to better understand the emergence and transmission of this novel clade in Bangladesh.

## MATERIALS AND METHODS

### Isolate Selection

From August-September 2021, as part of ongoing hospital surveillance, we identified a total of 60 *Candida auris* isolates and archived these isolates at the International Centre for Diarrhoeal Disease Research, Bangladesh (icddr,b) Mycology Laboratory. Isolates were cultured from skin swab samples from both axilla and groin for screening the patients admitted in the ICU and NICU of two tertiary care hospitals (one public and one private). In addition to on-admission screening sample, subsequent follow up samples were collected-after 72 hours of admission and before shifting from ICU-from patients to identify nosocomial colonization. [32]. Isolates from ICU patients were termed as BD-ICU, and isolates from NICU patients were termed BD-NICU throughout the manuscript.

### DNA Extraction and Whole Genome Sequencing

Ten *C. auris* isolates were randomly selected and retrieved from −80°C storage and grown on CHROMagar^TM^ Candida Plus media for presumptive identification of *C. auris*. Pure *C. auris* colonies were transferred from CHROMagar^TM^ plate to SDA plate and incubated at 37°*C* for 72 hours. Genomic DNA of the *C. auris* isolates was extracted using the Qiagen DNAeasy Blood & Tissue Kit, and the Illumina DNA Prep kit (Illumina Inc., San Diego, California, USA) was used for the construction of DNA libraries. Sequencing was performed on Illumina NextSeq 550 platform with 2×150 bp cycles at icddr,b Genome Centre.

### Variant Calling and Phylogenetic Analysis

To investigate the origin and possible route of transmission of sequenced *C. auris* isolates from Bangladesh, phylogenetic analysis was performed using the whole-genome sequences. A total of 25 isolates representative of *C. auris* clades I – V were also included in this analysis (**File S2**). Quality control, single nucleotide polymorphism (SNP) calling, and phylogenetic analysis were performed using the reference-based pipeline, MycoSNP, v1.4 (https://github.com/CDCgov/mycosnp-nf) with default parameters and using assembly B11205 (Accession no. GCA_016772135.1) generated from an isolate from India [33]. In brief, the MycoSNP workflow masks repetitive regions and indexes the reference genome in preparation for read alignment and snp calling. Paired-end reads were trimmed and filtered for low quality data. Samples, with at least 20X coverage and a % GC content between 42-47.5%, were used for alignment to the reference genome using the BWA [34] and pre-processed for downstream single-nucleotide polymorphism (SNP) analysis using GATK[35]. GATK *HaplotypeCaller* function is used for haplotype calling the processed BAM files from each isolate. The ploidy parameter for *C. auris* is set to haploid. *HaplotypeCaller* uses the PairHMM (Pair Hidden Markov Models) algorithm and performs pairwise alignment. Then, the resulting individual gVCF files are merged into a single multi-sample gVCF file by *CombineGVCFs* function. Next, the *GenotypeGVCFs* function performs SNP calling from this single gVCF file which results in a VCF file with all samples genotyped. GATK’s *VariantFiltration* function was used to filter SNP sites based on the QualityByDepth (QD), FisherStrand Score (FS) and RMSMappingQuality (MQ) parameters using filtering expression: “QD < 2.0 || FS > 60.0 || MQ < 40.0”. Additional customized SNP filters were applied based on parameters such as Genotype Quality (GQ), allele depth (AD), and depth (DP) using following criterion: : GQ ≥ 50, AD ≥ 80%, DP ≥ 10. The MycoSNP workflow recovered all SNP alleles from the VCF files using a Python script “vcfSnpsToFasta.py” (https://github.com/broadinstitute/broad-fungalgroup/tree/master/scripts/SNPs) to create an SNP alignment in multi-fasta format [33]. To construct Maximum Likelihood (ML) phylogenetic trees from SNP alignment, RAxML v8.2.12 [36] was utilized with the Generalized Time-Reversible model and a Gamma distribution (GTRGAMMA) to account for site-specific rate variation. Support for the ML phylogenetic trees were evaluated through 1000 bootstrap replicates of the alignment. The resulting trees were visualized using iTol v5.5 [37].

### Mutational Characterization

*De novo* assembly was performed using SPAdes v.3.15.5 to generate assemblies and assessed with Quast v.5.5.0 [38, 39]. The assembly reports were summarized in **File S5** in the supplemental material. A custom python script was generated to map *ERG11* (F126L, Y132F, K143R, and F444L), *CDR1* (V704L), *TAC1B* (S611P) and *MRR1A* (N647T) for azole resistance, *FCY1* (S70R) and *FUR1* (1Δ33, and F211I) for 5-flucytocine resistance and *FKS1* (S639Y/P/F, and F635C) genes for echinocandin resistance. Since the molecular mechanism of amphotericin B is not well established, we also investigated known mutations in *ERG3* (S58T) and *ERG6* (G403T, and R97fs) genes involved in ergosterol biosynthetic pathways which are postulated to confer amphotericin B resistance [14, 40].

### Antifungal Susceptibility Testing (AFST)

To assess the resistance of *C. auris* to antifungals, antifungal susceptibility testing (AFST) was performed using Gradient Diffusion Strips method or E-test (OFD-500-P03: https://www.cdc.gov/fungal/lab-professionals/pdf/afst-yeasts-h.pdf) according to Clinical and Laboratory Standards Institute (CLSI) guidelines [41]. The E-test gradient strips of fluconazole, voriconazole, caspofungin, and amphotericin B was obtained from Biomerieux. The reference number and lot number of corresponding anti-fungal strip are provided in **File S6**. The fluconazole concentration gradient ranged from 256 to 0.016 μg/ml while for other antifungals, it ranged from 32 to 0.002 μg/ml. The antifungals used in this study for AFST are representative of 3 major classes of antifungals (azoles, echinocandins and polyenes). The culture media used were Sabouraud dextrose agar (SDA; Becton Dickinson) and Roswell Park Memorial Institute medium 1640 (RPMI-1640; Sigma–Aldrich). Currently, there are no established minimum inhibition concentration (MIC) breakpoints for *C. auris.* CDC proposed tentative breakpoints for fluconazole (≥ 32 mg/L), voriconazole (≥ 1mg/L), caspofungin (≥ 2 mg/L), anidulafungin (≥ 4 mg/L), micafungin (≥ 4 mg/L), amphotericin B (≥ 2 mg/L) and other antifungals [7, 18, 42].

The *C. auris* isolates from this study were also tested for susceptibility to 5-flucytosine by disk diffusion method outlined by CLSI-M60 guidelines [43]. The assays were performed using fungal suspension adjusted to match the turbidity of a 0.5 McFarland standard. The antifungal disk for Flucytosine (AFY) 10 µg was purchased from Liofilchem^®^, Italy). Mueller Hinton II Agar (MHA; BD), Sabouraud Dextrose Agar (SDA; BD) and Roswell Park Memorial Institute Medium 1640 (RPMI-1640; Sigma–Aldrich) was used for preparation of media of the *C. auris* isolates. The reference and lot number of corresponding agar and media are provided in **File S6**. The disks were placed on to the inoculum media and the plates were incubated at 35°*C*(±2°*C*) for 18-24 hours. After 24 hours, the plates were examined and the zones of inhibition diameter (ZOD) around the discs were measured in millimeters (mm). The inhibition zone diameters (mm) were interpreted as Susceptible (S), Intermediate (I) and Resistant (R) based on established clinical breakpoints provided by CLSI. Susceptibility to 5-flucytosine was also tested via automated VITEK^®^2 system using an AST-YS09 card as per the manufacturer’s instructions. The time and temperature of incubation was similar to disk diffusion assay (18 − 24 *hours at* 35±2°*C*) but the results were expressed as MICs. The MIC calling range was 1 to 64 mg/L for 5-flucytosine. Since the VITEK^®^2 system lacks species-specific MICs for *C. auris*, the species was modified to different *Candida* spp. to obtain MICs for *C. auris* isolates. Notably, the MIC values remained consistent irrespective of the species assigned when retrieving MIC data for *C. auris* across all drugs. As *C. duobushaemulonii* closely related to *C. auris* and belongs to the *C. haemulonii* complex, analysis was based on MICs retrieved from this species [44, 45].

## Ethical approval

The study was approved by icddr,b Research Review Committee and Ethical Review Committee. The approval protocol number was PR-20122. All patients participated in this study gave written informed consent for donating the samples.

## Data availability

The sequencing data sets generated during this study are available in the Sequence Read Archive repository under projects PRJNA981511.

## ACKNOWLEDGMENTS

The study was funded by Centre for Disease Control and Prevention, Atlanta, USA, through cooperative agreement. We also gratefully acknowledge icddr,b core donors (Govt. of Bangladesh and Canada) for their unrestricted support and commitment to icddr,b’s research efforts.

Conceptualization, T.K., N.I.F., M.J., F.C., and M.R.; data curation, T.K., N.I.F., M.M.H., S.M., A.H., and D.S.; formal analysis, T.K., N.I.F., M.M.H., A.H., S.B.M., D.S., and L.A.P.; investigation, T.K., N.I.F., S.M., DA, M.J., N.A.C., F.C., and M.R.; methodology, T.K., N.I.F., M.M.H., A.H., S.B.M., DA, L.A.P., and M.J.; validation, T.K., N.I.F., A.H., S.B.M., D.S., DA, L.A.P., M.J., F.C., and M.R.; supervision, F.C., and M.R.; visualization, T.K., M.M.H., L.A.P., and N.A.C.; writing - original draft, T.K., N.I.F., and M.R.; writing -review & editing, T.K., N.I.F., M.M.H., S.M., A.H., S.B.M., D.S., DA, L.A.P., M.J., N.A.C., F.C., and M.R.

## Disclaimer

The findings and conclusions of this report are those of the authors and do not necessarily represent the official position of the Centers for Disease Control (CDC). The Sample IDs and Sequencing IDs used in this manuscript were randomly assigned, did not correspond to any of the hospital generated original IDs and were not disclosed to anyone outside the research group, not even to hospital staff or patients themselves.

## Supplemental Material

**Fig S1:** Phenotypic differences observed between BD Clade I and Clade VI isolates of *Candida auris* at different timelines (A) 24 hours (B) 48 hours and (C) 72 hours.

**File S1:** Patient metadata of BD *Candida auris* isolates.

**File S2:** *Candida auris* isolates used in this representing known five clades.

**File S3:** Quality metrices of BD *Candida auris* isolates generated by MycoSNP v1.4.

**File S4:** SNP distances generated by MycoSNP v1.4 for all *Candida auris* isolates in this study.

**File S5:** Assembly metrices of BD *Candida auris* isolates.

**File S6:** Anti-fungal strips, agar, and media used in this study with reference number and lot number.

