## Supplementary figures and images for "Emergence of the Novel Sixth *Candida auris* Clade VI in Bangladesh"

### Supplemental Figure 1

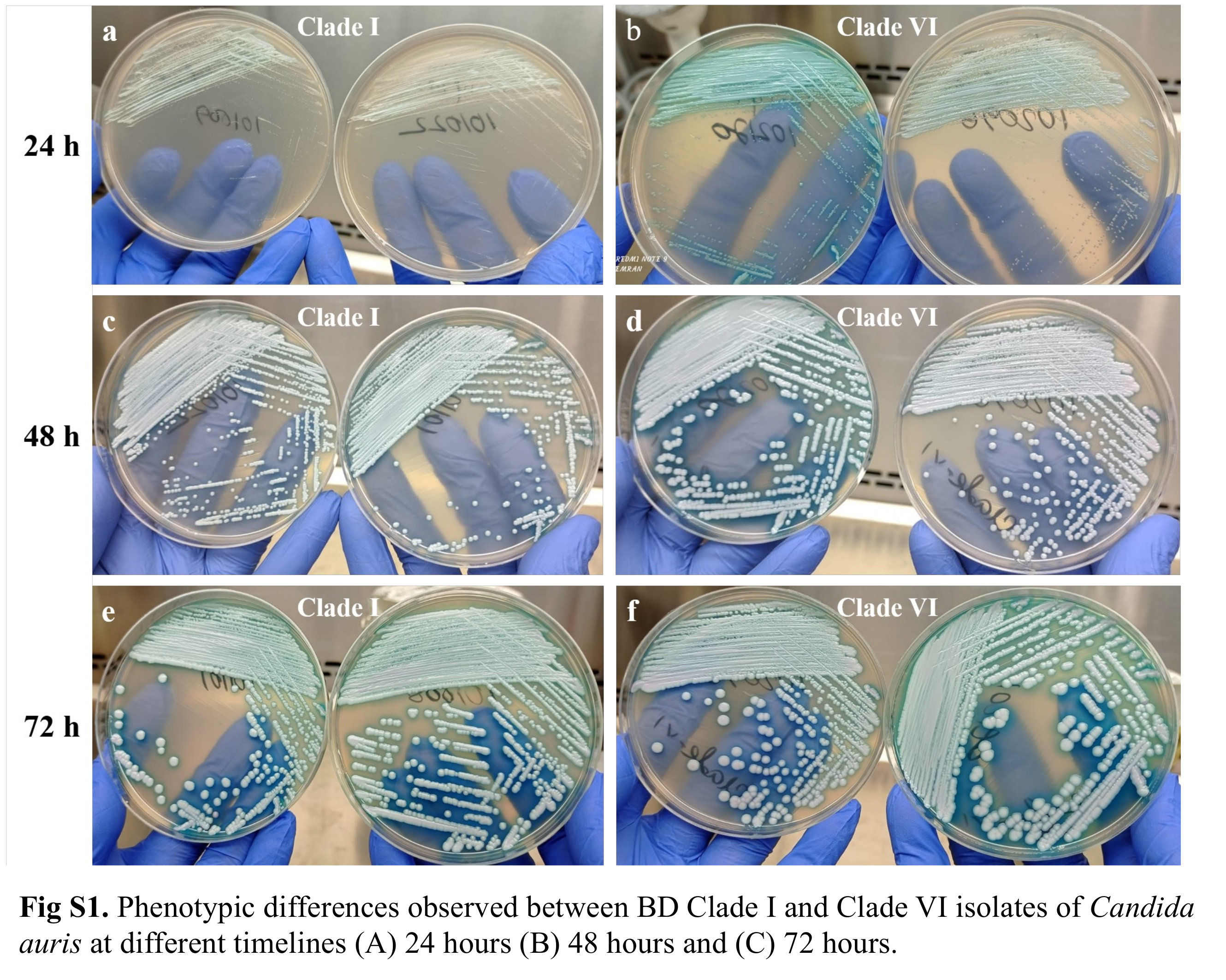
